# Characterizing the Clinical and Genetic Landscape of *KCNT1*-Related Disorders

**DOI:** 10.64898/2026.05.25.26354015

**Authors:** Sachi Lele, Ian McSalley, Shiva Ganesan, Alicia Harrison, Jan H. Magielski, Sarah M. Ruggiero, Anna J. Prentice, Nasha Fitter, Elise Brimble, Justin West, Mark P. Fitzgerald, Ingo Helbig, Jillian L. McKee

**Affiliations:** Division of Neurology, Children’s Hospital of Philadelphia, Philadelphia, PA, 19104, USA; The Epilepsy NeuroGenetics Initiative (ENGIN), Children’s Hospital of Philadelphia, Philadelphia, PA, 19104, USA; Department of Biomedical and Health Informatics (DBHi), Children’s Hospital of Philadelphia, Philadelphia, PA, 19146, USA; Department of Neurology, University of Pennsylvania Perelman School of Medicine, Philadelphia, PA, 19104, USA; Citizen Health, San Francisco, CA; KCNT1 Epilepsy Foundation, Scottsdale, AZ

**Keywords:** KCNT1, epilepsy, genetics, developmental and epileptic encephalopathy

## Abstract

*KCNT1*-related disorders represent clinically heterogeneous severe epilepsies associated with profound neurodevelopmental impairment. The full phenotypic spectrum and longitudinal disease trajectory remain incompletely characterized, which is a critical gap limiting the establishment of quantifiable endpoints necessary for future clinical trials. Compounding this challenge, identical pathogenic variants result in phenotypically distinct syndromes, including early infantile developmental and epileptic encephalopathy (EIDEE) and autosomal dominant sleep-related hypermotor epilepsy (ADSHE), underscoring unresolved genotype-phenotype relationships. To address these gaps, we performed a comprehensive analysis of 159 individuals with *KCNT1*-related disorders, including a longitudinally characterized subgroup of 62 individuals across 390 patient years, systematically defining disease progression, seizure trajectories, developmental outcomes, and treatment response across the full spectrum of the disorder. Seizures were nearly universal, affecting 157 of 159 individuals, with 81% (*n*=126/156) having seizure onset within the first year of life. Stratification by clinical subgroup revealed divergent seizure onset patterns. Recurrent variants did not significantly differ in age of seizure onset yet exhibited variant-specific clinical fingerprints, such as the preponderance of focal clonic seizures (OR=5.03, 95% CI 1.60-15.7, *f*=0.47) in those with the p.Gly288Ser variant. Comparison with a broader cohort of 14,893 individuals with neurodevelopmental disorders revealed phenotypic features such as migrating focal seizures (OR=21716, 95% CI 2409-Inf, *f*=0.42) and hypertonia (OR=26.5, 95% CI 18.2-38.3, *f*=0.45) to be more common in EIDEE, and nocturnal seizures (OR=29787, 95% CI 3062-Inf, *f*=0.5) and hyperactivity (OR=13.7, 95% CI 4.70-35.9, *f*=0.32) to be more common in ADSHE. These findings corroborate and extend those reported in the existing literature. Developmental milestones revealed marked delays across all domains. Analysis of longitudinal medication prescription patterns exposed striking therapeutic variability, reflecting the absence of a consistent treatment framework. Several anti-seizure medications frequently cited as beneficial, quinidine and cannabidiol, were not associated with seizure improvement or sustained seizure freedom in our cohort. In contrast, clobazam (OR=1.39, 95% CI 1.12-1.72, *f*=0.85), ketogenic diet (OR=1.30, 95% CI 1.07-1.57, *f*=0.75), and lacosamide (OR=2.03, 95% CI 1.54-2.66, *f*=0.59) demonstrated positive comparative effectiveness. Quantitative EEG analysis distinguished individuals with *KCNT1*-related disorders from age-matched controls with high accuracy (AUC=0.906), with key discriminating spectral features, including alpha power in the central and parietal regions, demonstrating significant reduction across childhood and adolescence. Collectively, these findings expand the phenotypic and genotypic landscape of *KCNT1*-related disorders through large-scale real-world clinical data, establish quantifiable longitudinal clinical endpoints, and provide actionable insights into genotype-phenotype relationships and differential treatment response. Together, these findings will help identify outcome measures and biomarkers to inform future clinical trial design.

## Introduction

Since the first pathogenic variant was identified in 2012, *KCNT1*-related epilepsies have remained one of the most severe and treatment-refractory developmental and epileptic encephalopathies (DEE).^1^ Frequent seizures and profound developmental delays define the condition, yet despite targeted therapies on the horizon, the true spectrum of disease burden and the metrics that best capture treatment response remain poorly understood. As clinical trials for genetically targeted therapies move closer to reality, establishing reliable, measurable endpoints has never been more urgent.

*KCNT1* encodes a sodium-activated potassium channel, with rare variants driving a clinically diverse spectrum of epilepsy and neurodevelopmental impairments. At one extreme sits epilepsy of infancy with migrating focal seizures (EIMFS), a devasting developmental and epileptic encephalopathy that falls within the broader category of early infantile developmental and epileptic encephalopathy (EIDEE), often leaving children with severe lifelong disabilities.^2^ At the other end, autosomal dominant sleep-related hypermotor epilepsy (ADSHE) presents with nocturnal seizures of later onset and, strikingly, largely intact development.^2^ Beyond these anchoring phenotypes, rare *KCNT1* variants have also been implicated in multifocal epilepsy and cardiac disturbances.^3^ Yet despite this breadth, clear genotype-phenotype correlations remain elusive.

Recurrent *KCNT1* variants can produce a range of different outcomes. Prior research has shown that the same mutation can manifest as either EIDEE or ADSHE, highlighting significant phenotypic variability.^3–5^ This phenotypic variability is more than a biological curiosity: it directly undermines efforts to standardize treatment, since medication responses are highly presentation-specific and, too often, limited. Therapeutic interventions, including quinidine treatment and small-molecule inhibitors, have been trialed, though their efficacy remains uncertain.^6,7^ Fully characterizing the clinical spectrum of *KCNT1*-related disorders is therefore not an academic exercise but a prerequisite for identifying clinically meaningful subgroups that may inform therapeutic development. While extensive prior work has highlighted key features of the disorder, including the early onset and high burden of seizures and developmental impairments, a more granular understanding of the natural history and key outcomes will be helpful for tailoring treatment decisions, guiding counseling, and designing future clinical trials.^1–3,5,6,8,9,23^

Electronic medical records (EMRs) offer a powerful, underutilized resource for capturing longitudinal clinical histories alongside genetic data.^10^ EMRs enable systematic tracking of how disease trajectories evolve over time through seizure type and frequency, treatment response, phenotypic features, and developmental trajectories. For this study, we combined data from the local EMR of a quaternary pediatric medical center, Citizen Health, a EMR data aggregator, and the previously published literature to assess key features of the disorder, including seizure trajectories, EEG findings, medication histories, developmental assessments, genetic reports, and general clinical features. Together, these data streams allow for a multidimensional characterization of disease progression and forms the basis for comprehensive phenotyping of *KCNT1*-related disorders.

Critically, this work is grounded in real-world data spanning many years of clinical follow-up, reflecting how patients present, are treated, and progress in practice. While numerous studies since 2012 have catalogued phenotypic features of *KCNT1*-related disorders in-depth, our goal was to track the full longitudinal clinical course including seizure frequency and severity changes, changes in treatment responsiveness, and achievement of developmental milestones.^1–3,5,6,8,9,23^ This work takes a first step towards expanding genotype-phenotype correlations, characterizing real-world treatment response patterns, and lays the groundwork for a natural history framework that can directly inform the design of future clinical trials and the identification of reliable, meaningful endpoints.

## Methods

### Cohort Overview and Inclusion Criteria

To identify individuals with *KCNT1*-related disorders, we conducted a systematic literature search of PubMed using “*KCNT1*” as the primary search term, retrieving existing published case reports and case series.^5,11–25^ Following review, a total of 97 previously reported individuals were identified for inclusion. We also incorporated individuals with EMR-derived data containing relevant clinical and genetic features, including 23 individuals from the Children’s Hospital of Philadelphia (CHOP) and 39 individuals from Citizen Health (an EMR data aggregator), yielding a longitudinal cohort of 62 individuals and a combined cohort of 159 individuals overall. All identified variants were verified as pathogenic or likely pathogenic by a licensed genetic counselor using American College of Medical Genetics (ACMG) criteria, incorporating prior disease associations from Human Gene Mutation Database (HGMD) and the published literature where available.^26,27^ To minimize duplication across sources, we performed systematic cross-referencing of literature-reported and EMR-derived records using variant, age of seizure onset, phenotypic syndrome, sex, and birth year as matching criteria. In instances where a given individual appeared to be represented in multiple sources, priority was assigned hierarchically: longitudinal EMR-derived data over literature-reported data, with CHOP records taking precedence over Citizen Health records in cases of overlap between the two EMR sources (*n*=4).

### Human Phenotype Ontology (HPO) Mapping of Phenotypic Features

Phenotypic features extracted from both EMR-derived data and the literature were systematically mapped to the Human Phenotype Ontology (HPO).^28–31^ Corresponding HPO identifiers were assigned to each feature to standardize terminology across sources and ensure consistent representation of clinical phenotypes. For data derived from Citizen Health, reported phenotypic descriptors were harmonized through conversion to their nearest equivalent standardized HPO terms.^28–31^ All annotated terms were subsequently subjected to automated ontological reasoning via term propagation within the HPO hierarchy, as done previously by our group, whereby specific phenotypic annotations were propagated to their corresponding ancestor terms, enabling comparison across the cohort even when clinical features are recorded at different levels of specificity in the medical records.^28–35^

### Mapping Phenotypes to a Common Framework

For each individual in our longitudinal cohort, clinical histories were reconstructed through systematic extraction of longitudinal data from EMRs on a month-by-month basis, encompassing seizure types and frequency, general clinical features, medication prescriptions, and developmental milestones, as previously done by our group.^32–35^ For individuals identified from the literature, available genetic findings and reported general clinical features were recorded. The epilepsy syndrome classification, corresponding variant data, and age of seizure onset were documented in all individuals. Phenotypic features were mapped to HPO terms as described above. Seizure frequency (SF) was recorded utilizing the scale from the Pediatric Epilepsy Learning Health Systems (PELHS) as follows: multiple daily seizures (>5 per day, SF score = 5), several daily seizures (2–5 per day, SF score = 4), daily seizures (SF score = 3), weekly seizures (SF score = 2), monthly seizures (SF score = 1) and no seizures (SF score = 0).^32–36^

### Anti-Seizure Medication Histories and Comparative Effectiveness

For each individual in the longitudinal cohort, anti-seizure medication (ASM) exposure was systematically reconstructed by delineating the first prescribed month and last prescribed month of each agent, with any intervening discontinuation and re-initiation explicitly captured. A total of 18 ASMs were included in this analysis. Rescue medications were excluded, while the ketogenic diet and hormonal therapy were incorporated as therapeutic interventions of interest. Comparative effectiveness was assessed by evaluating seizure frequency during periods concurrent with each ASM prescription relative to non-concurrent periods, as previously described.^32–35^ Two composite clinical outcomes were defined: seizure improvement and seizure freedom for some period of time. Seizure improvement was defined as a reduction in seizure frequency of at least one interval on the PELHS scale; for example, a decrease from multiple daily seizures (>5 per day, SF = 5) to daily seizures (SF = 3).^32–26^ Seizure freedom was defined as the transition from any seizure activity to complete seizure cessation (SF = 0).^32–36^ Comparative effectiveness across ASMs was evaluated using odds ratios derived from Fisher’s exact test, with both clinical outcomes combined as the primary endpoint.

### Data Analysis and Statistical Testing

In order to illustrate the unique characteristics of *KCNT1*-related disorders, phenotypic features from 14,893 individuals with neurodevelopmental disorders were aggregated from two previously published cohorts and compared with those observed in our *KCNT1* cohort.^37,38^ Odds ratios were calculated using Fisher’s exact test to quantify the relative association of each feature with *KCNT1*. Comparative analyses were additionally performed within our *KCNT1* cohort across clinical subgroups and recurrent variants, with odds ratios used to assess feature enrichment across groups. The progression of developmental milestones achieved was compared to the Centers for Disease Control and Prevention’s (CDC) typical development timeline.^39^ Statistical testing for association and similarity analyses were performed with correction for multiple comparisons using a 5% False Discovery Rate (FDR).

### qEEG Methods

EEGs obtained through routine clinical care and recorded using the standard 10-20 system of electrode placement for individuals with *KCNT1* were extracted from the CHOP clinical EEG database (Neuroworks) and analyzed through a custom-built Python pipeline as previously described.^40^ In brief, the pipeline uses automated independent component analysis to remove sections of EEG with eye movement, cardiac, and muscle artifacts. EEG segments annotated as sleep, seizures, or activating procedures were also excluded. Only EEGs with at least 15 clean 4-second epochs were selected for further analysis. EEG spectral bandpower was estimated using the Welch method across standard frequency bands, and relative bandpower and frequency band ratios were calculated. We selected age-matched controls from our previously described control cohort and then trained random forest models to classify EEGs as originating from individuals with *KCNT1* or neurotypical controls.^40^ Then we evaluated the spectral features with the highest Gini importance as potential biomarkers of *KCNT1*-related epilepsy.

## Results

### The variant landscape of *KCNT1* is characterized by recurrent missense variants

*KCNT1-*related disorders are notable for clinical heterogeneity, in which the same pathogenic variant can result in different clinical phenotypes, complicating genotype-phenotype correlations. Here, we assessed the clinical features of 159 individuals with *KCNT1*-related disorders, of whom 132 were classified as EIDEE and 22 as ADSHE. The remaining five individuals carried pathogenic variants but exhibited seizure and neurological features that did not align with either category. To evaluate genotype-phenotype associations, we first examined the spectrum of *KCNT1* variants identified across the cohort, which revealed several recurrent variants including p.Arg474His/Cys (*n*=23), p.Ala934Thr/Ser (*n*=20), p.Gly288Ser (*n*=19), and p.Arg398Gln/Leu (*n*=11), consistent with previously reported recurrent *KCNT1* variants in the literature (**Fig. 1A**).^41^ All identified variants were missense variants, a feature unique to *KCNT1* compared to other related epilepsy genes (**Fig. 1B**). Individuals with recurrent variants were categorized by clinical subgroup, with no clear genotype-subgroup correlations identified (**Fig. 1C**). While those with the p.Arg474His/Cys presented with EIDEE, those carrying p.Gly288Ser and p.Arg950Gln presented with either EIDEE or ADSHE. Two individuals with atypical developmental and epileptic encephalopathy carried either p.Ala934Thr/Ser or p.Arg398Gln/Leu. Together, these findings highlight the clinical and molecular heterogeneity of *KCNT1*-related disorders, underscoring the complexity of genotype-phenotype correlations.

**Figure 1.**
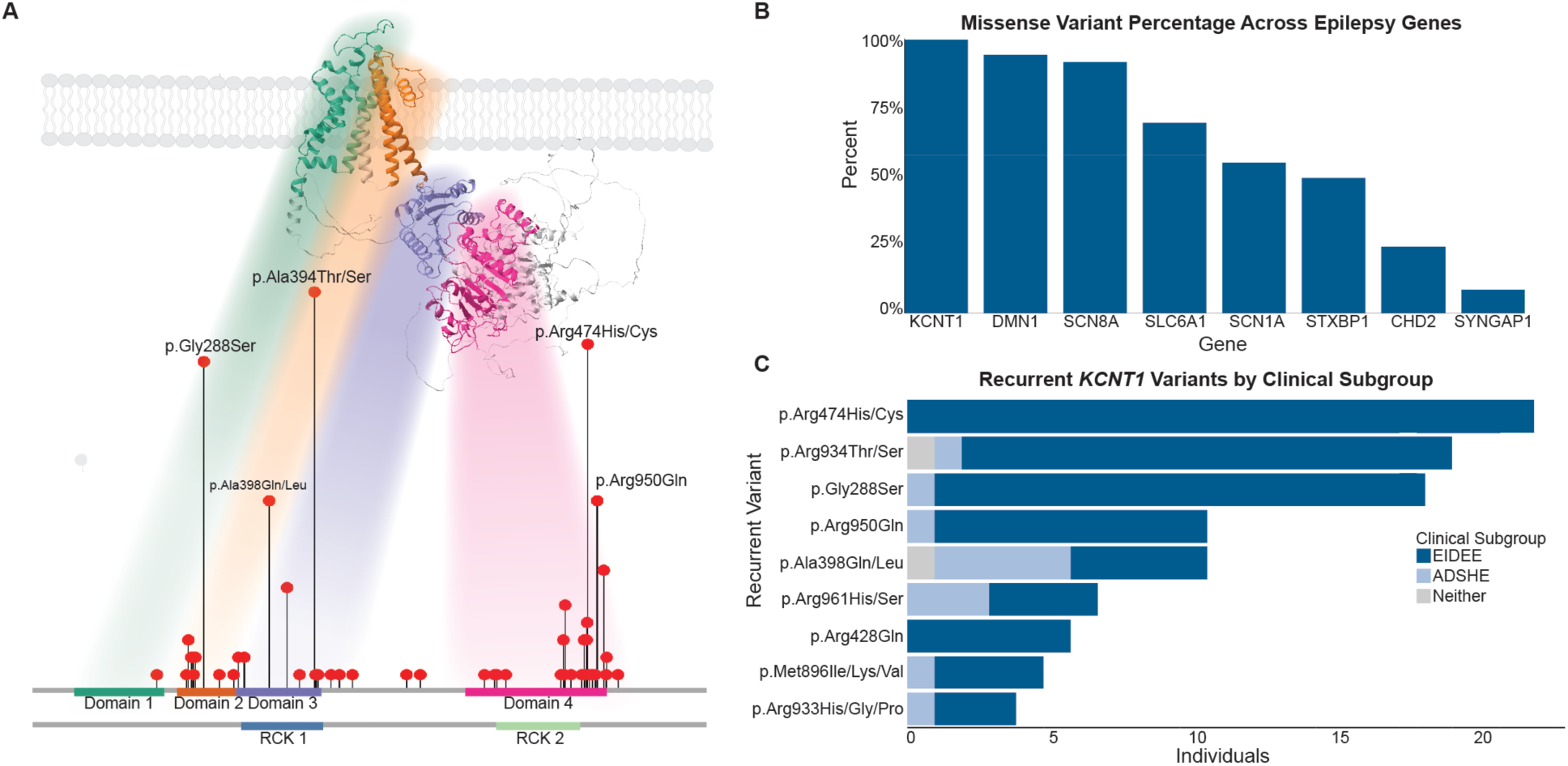
Overview of *KCNT1* variants. (**A**) Recurrent *KCNT1* variants mapped onto the protein structure, with transmembrane domains (Domains 1–4) and intracellular RCK domains highlighted. Colored gradients indicate the position of each domain in both the folded and extended visualizations. Labeled variants represent the most frequently observed pathogenic substitutions. (**B**) Percentage of missense variants in *KCNT1* compared to other epilepsy genes. (**C**) Distribution of recurrent *KCNT1* variants in our cohort, grouped by clinical subgroup (EIDEE, ADSHE, unknown classification).

### *KCNT1*-RD have a unique phenotypic signature with defined subgroups

Although broad phenotypic features have been previously described, the full clinical spectrum and variability across affected individuals with *KCNT1*-related disorders remain incompletely understood. We therefore examined clinical phenotypes in our cohort of 159 individuals, with the twenty most common features summarized in **Figure 2A**. Seizures were highly abundant (frequency (*f*)=0.98) and occurred across a range of seizure types, including motor (*f*=0.69), bilateral tonic-clonic (*f*=0.43), tonic (*f*=0.3), and nocturnal seizures (*f*=0.094). Musculoskeletal abnormalities (*f*=0.67) were also frequently observed, most commonly involving disturbances in muscle tone (*f*=0.61), including hypotonia (*f*=0.44) and hypertonia (*f*=0.39). Abnormalities of the respiratory (*f*=0.39) and digestive (*f*=0.41) systems were the most common non-neurological manifestations.

**Figure 2.**
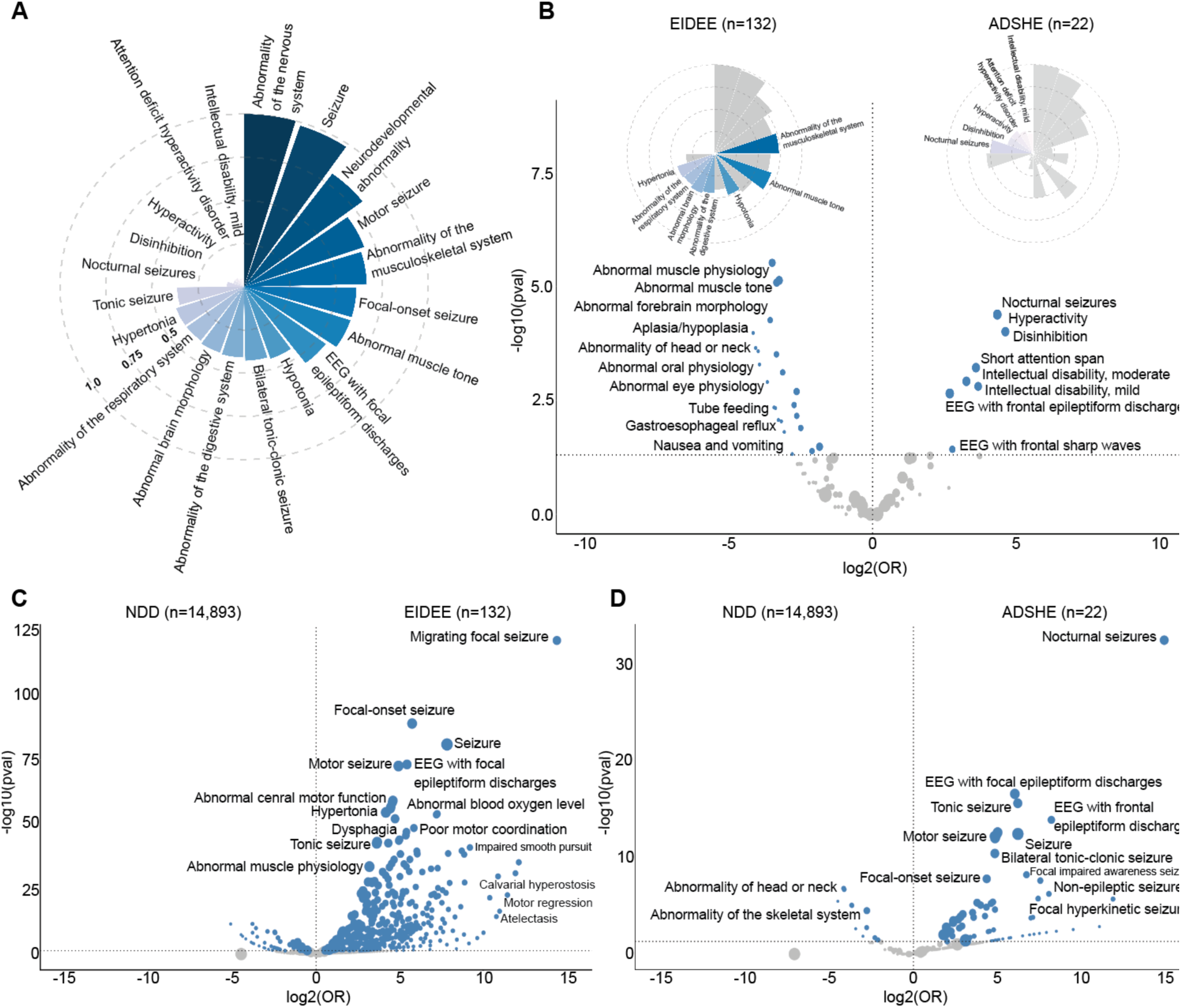
Common phenotypic features in *KCNT1*-related disorders. (**A**) The twenty most frequent clinical features across the entire cohort *(n*=159), with concentric rings indicating proportion of individuals affected. (**B**) Clinical features enriched in the EIDEE or ADSHE subgroups, with significant features from the twenty most common highlighted in their respective radar plots. (**C**) Features commonly associated with the EIDEE cohort (*n*=132) compared to a large reference cohort (*n*=14,893) of individuals with neurodevelopmental disorders. (**D**) Features commonly associated with the ADSHE cohort (*n=*22) compared to the same reference cohort. Blue dots and highlights indicate a *P-*value < 0.05.

Of note, major aortopulmonary collateral artery (MAPCA), a cardiovascular phenotype, was observed in seven individuals (*f*=0.044). Although MAPCA is not directly represented within the biomedical ontology used here, related features such as “aortopulmonary collateral arteries” captured through HPO terminology corresponded with the reported MAPCA phenotype.^24–26^ Of the seven, three individuals had explicitly documented MAPCA diagnoses, while the remaining four were inferred to have MAPCA-associated pathology based on cardiovascular phenotypes reported through Citizen clinical data. Ages of diagnoses ranged from 2 months (*n*=2), 4 months (*n*=2), 11 months (*n*=1), to 20 months (*n*=2), with this variability in age at diagnosis signifying the importance of early screening and intervention to mitigate severe complications associated with MAPCAs.^43=42^

To better contextualize variability within *KCNT1*-related disorders, we compared the two primary clinical subgroups, EIDEE and ADSHE, to delineate patterns of overlap and divergence across the phenotypic spectrum (**Fig. 2B**). This comparison revealed a clear distinction in the distribution of common features across these clinical subgroups. Individuals with EIDEE were more likely to exhibit severe neurologic and systemic impairments, including abnormalities of muscle tone (OR=9.3, 95% CI 2.77-39.8, *f*=0.69), spanning both hypotonia (OR=5.82, 95% CI 1.58-31.9 *f*=0.49) and hypertonia (OR=16.7, 95% CI 2.50-701.5, *f=*0.46), as well as a broader involvement of the musculoskeletal system (OR=12.0, 95% CI 3.53-51.5, *f=*0.74). Digestive system abnormalities were also common (OR=4.86, 95% CI 1.32-26.8, *f*=0.45), including feeding tube dependence (OR=10.1, 95% CI 1.50-427.5, *f=*0.34) and gastroesophageal reflux (OR=8.82, 95% CI 1.31-373.6, *f*=0.31). Additional features more common in the EIDEE group included respiratory system abnormalities (OR=15.7, 95% CI 2.35-660.9, *f*=0.44) and eye abnormalities (OR=5.3, 95% CI 1.19-48.5, *f=*0.36), further reinforcing the severe and multisystemic nature of this phenotype. Individuals with EIDEE also had 17-fold higher odds of abnormalities of brain morphology (OR=17.2, 95% CI 2.57-722.7, *f*=0.46), particularly morphological abnormalities of the forebrain (OR=15.7, 95% CI 2.35-660.9, *f=*0.44). In contrast, the ADSHE subgroup was characterized by a profile skewed toward behavioral and neuropsychiatric features. Individuals more commonly presented with disinhibition (OR=20.2, 95% CI 3.95-128.6, *f=*0.31), short attention span (OR=12.1, 95% CI 2.49-63.0, *f=*0.27), and hyperactivity (OR=20.2, 95% CI 3.95-128.6, *f*=0.32), as well as formal diagnoses of attention deficit hyperactivity disorder (OR=24.6, 95% CI 3.82-256.9, *f=*0.27). The odds of nocturnal seizures were 66 times higher in individuals with ADSHE (OR=65.5, 95% CI 11.5-631.3, *f*=0.5), and they had characteristic EEG findings with increased frontal epileptiform discharges (OR=6.43, 95% CI 1.73-22.5, *f*=0.32). The degree of intellectual disability in individuals with ADSHE was more commonly mild than in EIDEE (OR=12.7, 95% CI 2.19-86.8, *f*=0.23). Together, these findings define the phenotypic landscape of *KCNT1*-related disorders and reveal distinct clinical signatures across the EIDEE and ADSHE subgroups.

To deepen our understanding of the clinical features associated with *KCNT1*-related disorders, we compared phenotypes enriched across both clinical subgroups against a broader cohort of 14,893 individuals with neurodevelopmental disorders.^33,34^ Individuals with EIDEE showed enrichment for seizure-related phenotypes relative to the larger cohort. In fact, the odds of seizures were 228 times higher (OR=228.1, 95% CI 61.8-1922, *f*=0.98), with these individuals more likely to have focal-onset seizures (OR=53.7, 95% CI 36.7-79.2, *f*=0.65), and specifically migrating focal seizures (OR=21716, 95% CI 2409-Inf, *f*=0.42). Motor seizures (OR=30.50, 95% CI 20.7-45.5, *f*=0.70) were also significantly enriched, alongside characteristic features such as abnormal muscle physiology (OR=12.4, 95% CI 8.42-18.5 *f*=0.71), including hypertonia (OR=26.5, 95% CI 18.2-38.3, *f*=0.45), and gastroesophageal reflux (OR=8.36, 95% CI 5.57-12.3, *f*=0.30). Our ADSHE subgroup also showed a distinct phenotypic profile, with a dramatic overrepresentation of nocturnal seizures (OR=29787, 95% CI 3062-Inf, *f*=0.5), bilateral tonic-clonic seizures (OR=28.4, 95% CI 11.1-75.5, *f=*0.5), and focal hyperkinetic seizures (OR=3633, 95% CI 131.3-Inf, *f*=0.091). EEG findings were notable for a 64-fold higher odds of focal epileptiform discharges (OR=64.4, 95% CI 25.0-178.4, *f*=0.64). The ADSHE group was again characterized by clinical features including disinhibition (OR=7.60, 95% CI 2.76-19.5, *f*=0.36) and hyperactivity (OR=13.7, 95% CI 4.70-35.9, *f*=0.32). Thus, leveraging this larger cohort enables clearer delineation and comparison of the distinct phenotypic features associated with clinical subgroups in *KCNT1-*related disorders.

### Recurrent variants have unique phenotypic fingerprints

Identical pathogenic variants in *KCNT1* can give rise to distinct clinical subgroups, making the identification of variant-specific clinical features important for clarifying the association between genotype and phenotype. We therefore analyzed these relationships for four recurrent *KCNT1* variants in our cohort of 159 individuals (**Fig. 3**), identifying distinct phenotypic features associated with specific variants. Individuals carrying the recurrent p.Arg474His/Cys variant (*n*=23) exhibited a broad clinical profile marked by both neurological and systemic involvement. Neurologically, these individuals more frequently presented with global developmental delay (OR=4.35, 95% CI 1.59-12.3, *f*=0.57), cerebral visual impairment (OR=7.37, 95% CI 2.13-25.5, *f*=0.35), choreoathetosis (OR=9.80, 95% CI 1.06-124.2, *f*=0.13), and infantile spasms (OR=3.46, 95% CI 0.83-12.8, *f*=0.22). This variant was also associated with notable systemic manifestations, including respiratory distress (OR=4.09, 95% CI 1.20-13.1, *f*=0.30), heart murmur (OR=3.87, 95% CI 0.91-14.7, *f*=0.22), and feeding tube dependence (OR=5.20, 95% CI 1.89-15.1, *f*=0.61). Individuals with the p.Ala934Thr/Ser variant (*n*=20) demonstrated a phenotype characterized by difficulties with coordinated movements, including abnormal conjugate eye movements (OR=4.42, 95% CI 1.37-13.8, *f*=0.40), and oral-pharyngeal dysphagia (OR=6.87, 95% CI 1.71-26.6, *f*=0.30). The p.Gly288Ser variant (*n*=19) was largely defined by seizure-related manifestations, including focal clonic seizures (OR=5.03, 95% CI 1.60-15.7, *f*=0.47), but they also demonstrated a 6.6-fold higher odds of hypsarrhythmia (OR=6.65, 95% CI 1.46-28.3, *f*=0.26), 15.8-fold higher odds of Chiari type 1 malformation (OR=15.8, 95% CI 0.78-964.2, *f*=0.11), and 2.9-fold higher odds of severe intellectual disability (OR=2.90, 95% CI 0.95-8.71, *f*=0.47). Finally, individuals harboring the p.Arg398Gln/Leu variant (*n*=11) showed a distinct pattern centered on language development, with delays in both receptive (OR=25.7, 95% CI 2.58-347.5, *f*=0.27) and expressive (OR=17.2, 95% CI 2.0-151.6, *f*=0.27) language being prominent. Although variant-specific clinical features were identified, these features notably did not segregate clearly with either the EIDEE or ADSHE clinical subgroups, suggesting that these traditional broad categories may not capture all of the variability within the disorder.

**Figure 3.**
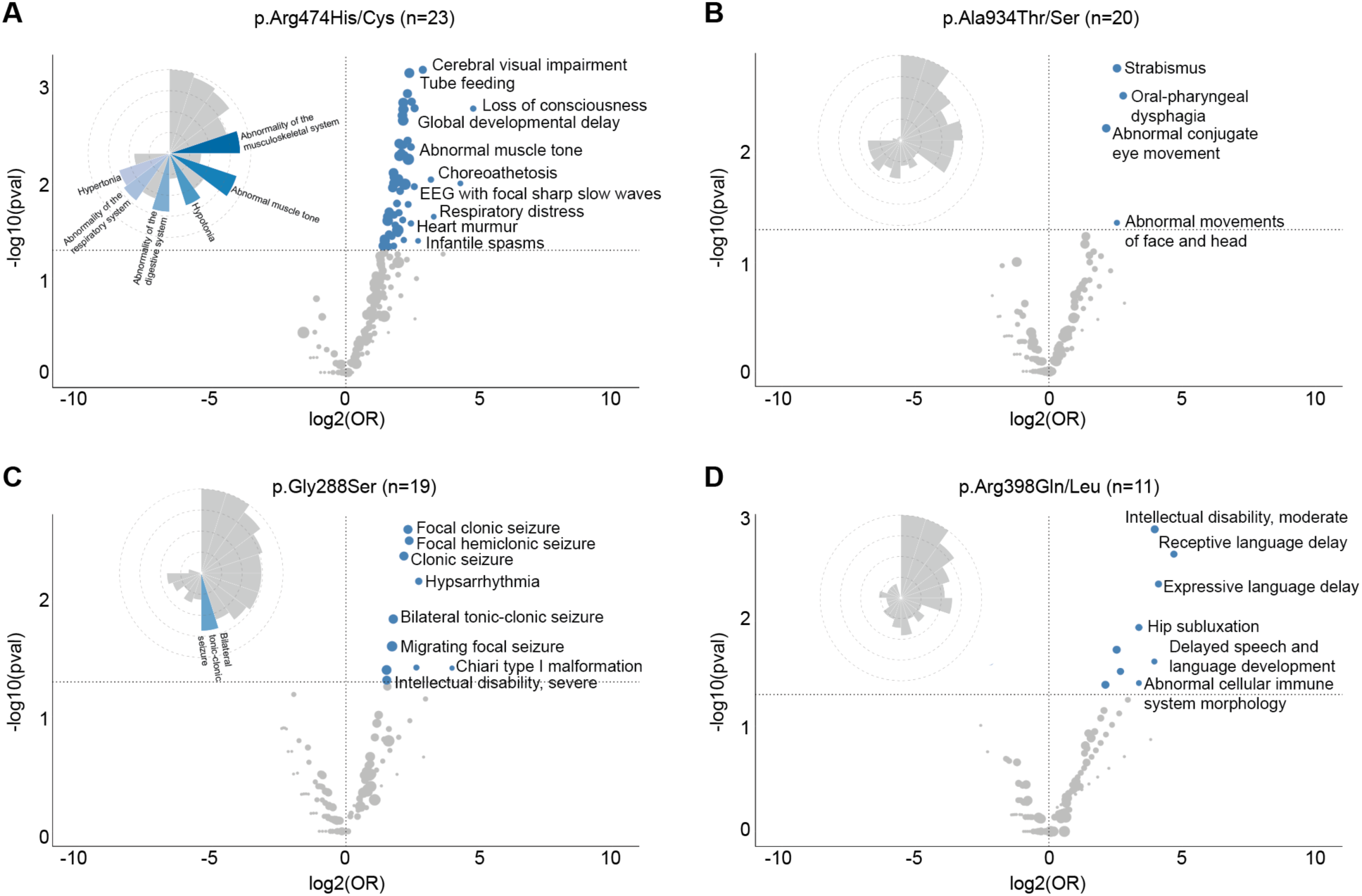
Genotype-phenotype correlations in *KCNT1* recurrent variants. Clinical features commonly associated with the (**A**) p.Arg474His/Cys, (**B**) p.Ala934Thr/Ser, (**C**) p.Gly288Ser, and (**D**) p.Ala398Gln/Leu recurrent variants, compared to the remainder of the cohort. Significantly enriched features from the twenty most common phenotypes (Figure 2A) are highlighted in the inset radar plots. Blue dots and highlights indicate p*-*values < 0.05.

### Longitudinal disease histories reveal early seizures and developmental delay

While we have characterized the genotype-phenotype correlations across recurrent variants and clinical subgroups, these analyses reflect static, cross-sectional snapshots of *KCNT1-*related disorders. To extend beyond this framework, we next examined how phenotypic features unfold over time to enable a dynamic view of disease progression. Across our cohort of 62 individuals, a total of 390 patient-years and 765 unique phenotypic terms were captured. The 100 most common features illustrate patterns of phenotypic emergence and persistence, demonstrating a high burden of disease early in life (**Fig. 4A**). Specifically, we see that seizure burden remained high at multiple timepoints, with the vast majority of individuals with at least daily seizures within any monthly time bin between birth and the age of 5 years, suggesting that seizures represent a sustained and defining feature of the disease course. In contrast, when assessing respiratory system abnormalities, the longitudinal data reveal a higher frequency observed early in life, with 37% (*n*=23/62) of the cohort affected at 18 months of age, which declines over time to 15% (*n*=9/62) affected at 5 years of life, indicating a more transient impact of respiratory symptoms. Taken together, these findings suggest that phenotypic features in *KCNT1*-related disorders follow distinct temporal trajectories that may inform the timing of clinical management and evaluation.

**Figure 4.**
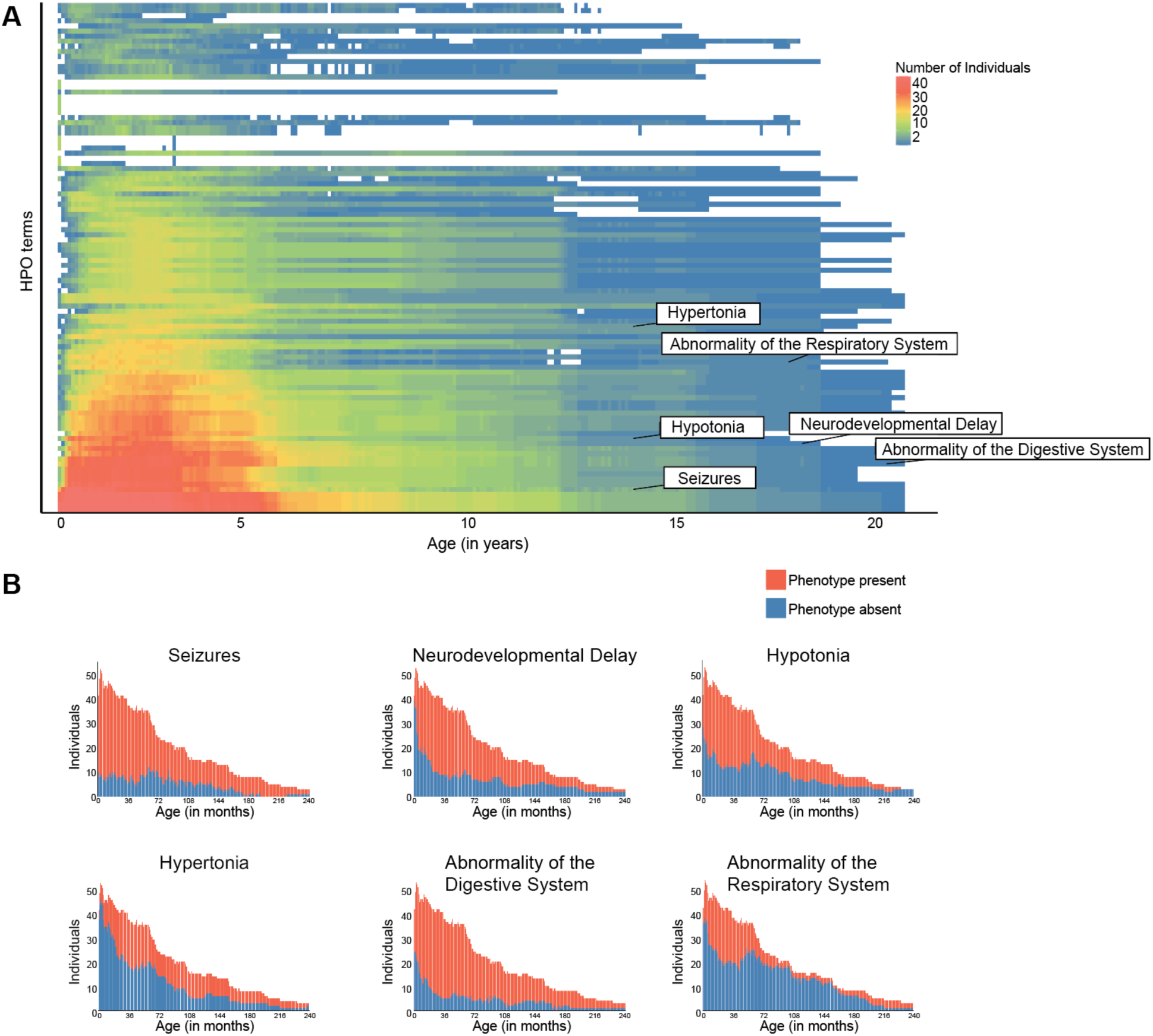
Longitudinal clinical histories in *KCNT1*. (**A**) Heatmap of the top 100 HPO terms across the cohort, ordered by number of individuals affected. Each row represents a clinical feature; color indicates the number of individuals presenting with that feature at each timepoint. Selected features of interest are labeled. (**B**) Clinical histories of six selected features displayed as stacked bar charts, binned by month. Red indicates the number of individuals with the feature present and blue indicates those without.

### *KCNT1*-related epilepsy is defined by early refractory seizures

Although seizures are a well-established clinical feature of individuals with *KCNT1*-related disorders, quantifying how seizure frequency evolves over time is important for defining meaningful clinical endpoints and assessing treatment response. Within the cohort of 159 individuals, two individuals never had seizures and a single individual did not have a reported seizure onset, resulting in an analyzable cohort of 156 out of 159 individuals. Overall, 81% (*n*=127/156) had seizure onset by one year of age. This trend was especially pronounced among individuals with EIDEE, of whom 93% (*n*=120/129) developed seizures before one year of age with a median seizure onset of one month, compared with 23% (*n=*5/22) of individuals with ADSHE having seizures in the first year of life with a median seizure onset of 66 months (**Fig. 5A**, *P*<0.001 for onset in EIDEE/ADSHE subgroups). In contrast, seizure onset did not differ among recurrent variants (*P*>0.05). 95% of individuals with the p.Arg474His/Cys (*n*=22/23) or p.Gly288Ser (*n*=18/19) variants, 90% (*n*=18/20) of individuals with the p.Ala934Thr/Ser variant, and 85% (*n*=9/11) of individuals with the p.Arg398Gln/Leu variant had seizures by one year of age.

**Figure 5.**
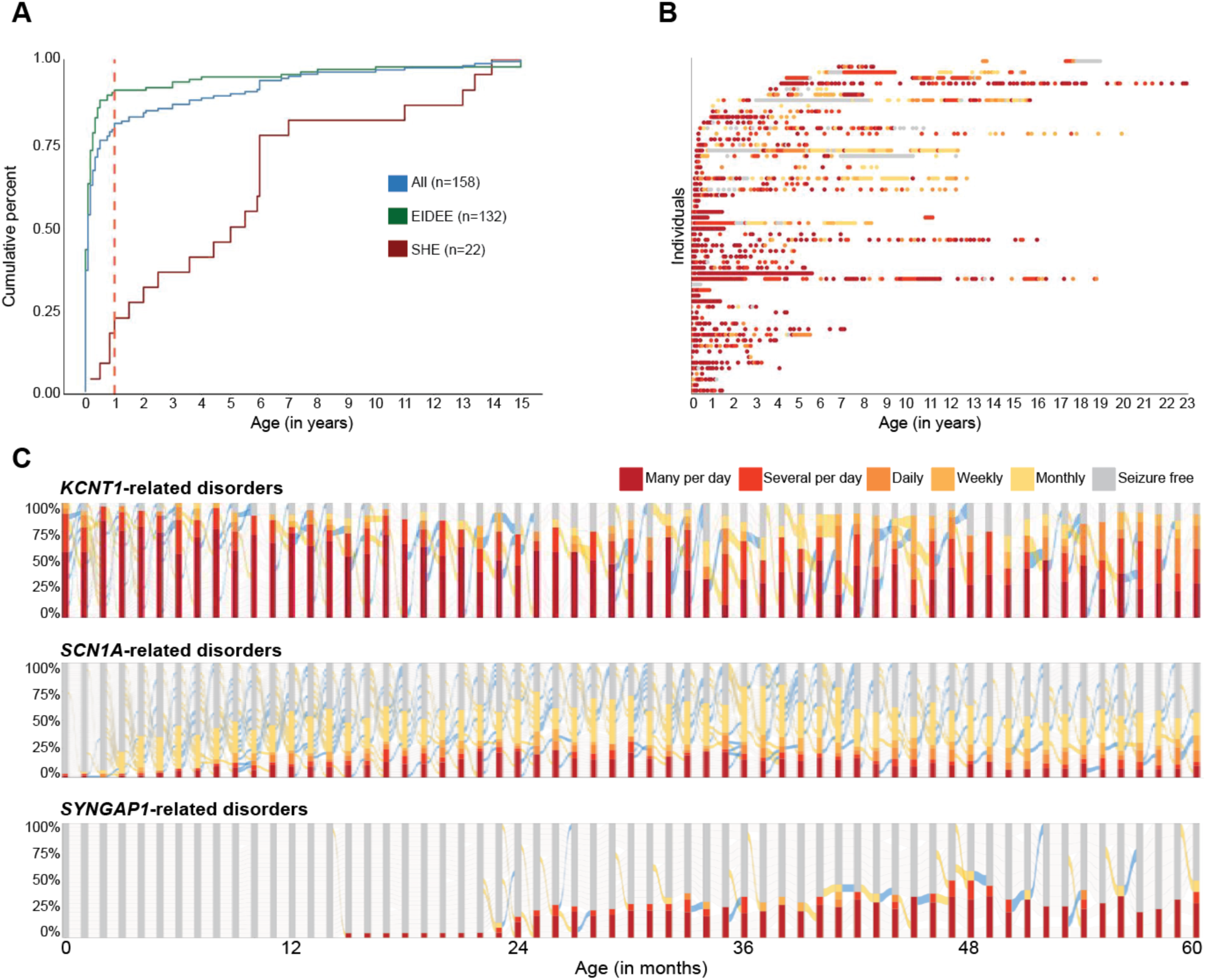
Seizure histories in *KCNT1*-related disorders. (**A**) Cumulative seizure onset across the entire cohort, subdivided by clinical subgroup. (**B**) Individual-level seizure histories ordered by seizure onset. Each dot represents a seizure frequency recorded in the EMR, coded by color: many per day (SF=5, dark red); several per day (SF=4, red); daily (SF=3, dark orange); weekly (SF=2, light orange); monthly (SF=1, yellow); seizure-free (SF=0, grey). (**C**) Changes in seizure frequency on a month-by-month basis, with the proportion of individuals at each severity level shown as stacked bars. Connecting lines indicate transitions in seizure frequency between consecutive months: blue indicates improvement, yellow indicates worsening, and grey indicates no change in seizure frequency. *KCNT1*-related disorders are shown alongside *SCN1A*- and *SYNGAP1*-related disorders for comparison.

Seizure frequency was tracked over time on a month-by-month basis using data from the EMR and Citizen Health in the longitudinal cohort of 62 individuals (**Fig. 5B**). All individuals developed seizures at some point in their lives, but two individuals did not have reported seizure frequencies and were not included in the analysis. Most individuals (*n*=55/60) had a high seizure burden, defined as at least five seizures per day, which frequently persisted throughout life. However, variability in seizure severity was observed, with some individuals (*n*=20/60) transitioning to weekly or monthly seizures later in life. Notably, only 10 out of 60 individuals had seizure freedom for more than one month at some point during follow-up. These findings highlight the persistent and severe nature of seizures in *KCNT1*-related disorders.

Of note, all 21 individuals with documented seizure information in the first month of life had seizures, with 57% (n=12/21) having at least five daily seizures, and the proportion of individuals with seizures only declined marginally by the age of five years. The early onset and severity of *KCNT1*-related epilepsy is further emphasized by comparisons with other genetic epilepsies (**Fig. 5C**). For example, individuals with *SCN1A*-related disorders typically have seizure onset within the first year of life, even though seizures tend to occur less frequently.^42^ In contrast, individuals with S*YNGAP1-*related disorders generally present with seizure onset after the first year of life.^35^ Together, these comparisons reinforce the early onset and sustained severity of seizures in *KCNT1*-related disorders.

### Developmental milestone acquisition defines measurable clinical trial endpoints

As with seizures, developmental delay is a prominent feature of *KCNT1*-related disorders; accordingly, delineating the timing of milestone acquisition within a large cohort may facilitate the understanding of the overall clinical progression. We assessed the time course of developmental milestone acquisition across the longitudinal cohort of 55 individuals with EIDEE, excluding four individuals with ADSHE, two individuals classified as atypical developmental and epileptic encephalopathy, and one individual with EIDEE without quantifiable developmental milestone acquisition (**Fig. 6A**). Many individuals (*n*=7/55) within the cohort were unable to reach any of the assessed milestones, emphasizing the severity of *KCNT1*-related disorders. Among individuals who achieved at least one of the milestones quantified in our analysis (*n*=48/55), milestones were typically achieved later in life compared to neurotypical developmental timelines and other neurogenetic conditions.^32,33,42,43^ For example, only 4 out of 31 (13%) individuals with documented verbal communication skills were able to attain this milestone, with 3 out of 4 (75%) achieving verbal communication by five years. Additionally, 6 out of 39 (15%) individuals attained walking, with 2 out of 6 (33%) achieving independent ambulation by 18 months. However, the remaining four individuals achieved this milestone later, indicating variability in motor development amongst individuals living with *KCNT1*-related disorders. Thus, these findings support the use of developmentally anchored and temporally defined outcomes as meaningful clinical trial endpoints.

**Figure 6.**
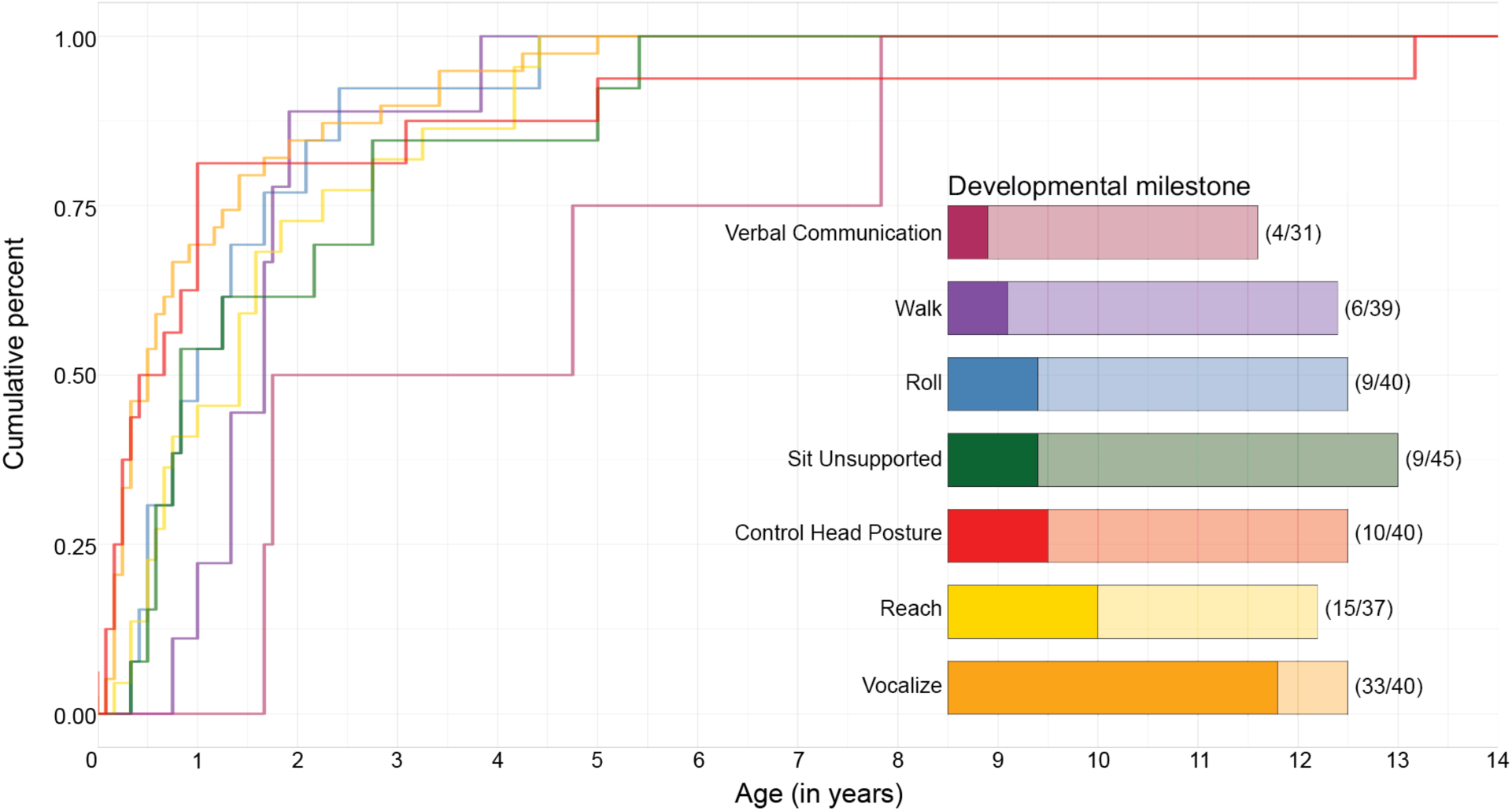
Developmental milestone acquisition in *KCNT1*. Cumulative proportion of individuals acquiring each developmental milestone over time, for those individuals who achieved that milestone. The inset bar graph indicates the number of individuals who acquired each milestone (dark shading) out of those assessed.

### *KCNT1*-related epilepsy necessitates a wide range of therapeutic strategies

Although several anti-seizure medications (ASMs) have been reported in the literature to improve seizure control, treatment responses remain variable.^6,44,45^ This is reflected in the persistent seizure burden observed in this cohort of 62 individuals. To characterize treatment patterns over time, we assessed the frequency of medication prescriptions across the lifespan, restricting inclusion to therapies prescribed concurrently in at least three individuals (**Fig. 7A**). One individual had listed medications but did not have corresponding monthly start and end dates, resulting in a cohort of 61 individuals. No single anti-seizure medication or treatment was consistently used across all individuals with *KCNT1*-related disorders. However, several therapies, including levetiracetam (*n*=58/61), clobazam (*n*=50/61), phenobarbital (*n*=47/61), ketogenic diet (*n*=44/61), and topiramate (*n*=41/61), were among the most frequently used. Among these, the ketogenic diet demonstrated the longest average duration of use (37.6 months), with sustained utilization suggesting potential efficacy. In contrast, levetiracetam (3.9 months), clobazam (5.7 months), phenobarbital (2.1 months), and topiramate (2.3 months) had relatively short average durations of use. Individuals with *KCNT1*-related disorders trialed a substantial number of medications overall, with a mean of 11.3 (median=11, range 1-25) unique medications per individual. The median age of medication initiation was 2 months (mean=11.2, range 0-196), highlighting the early onset and aggressive management of seizures in this population.

**Figure 7.**
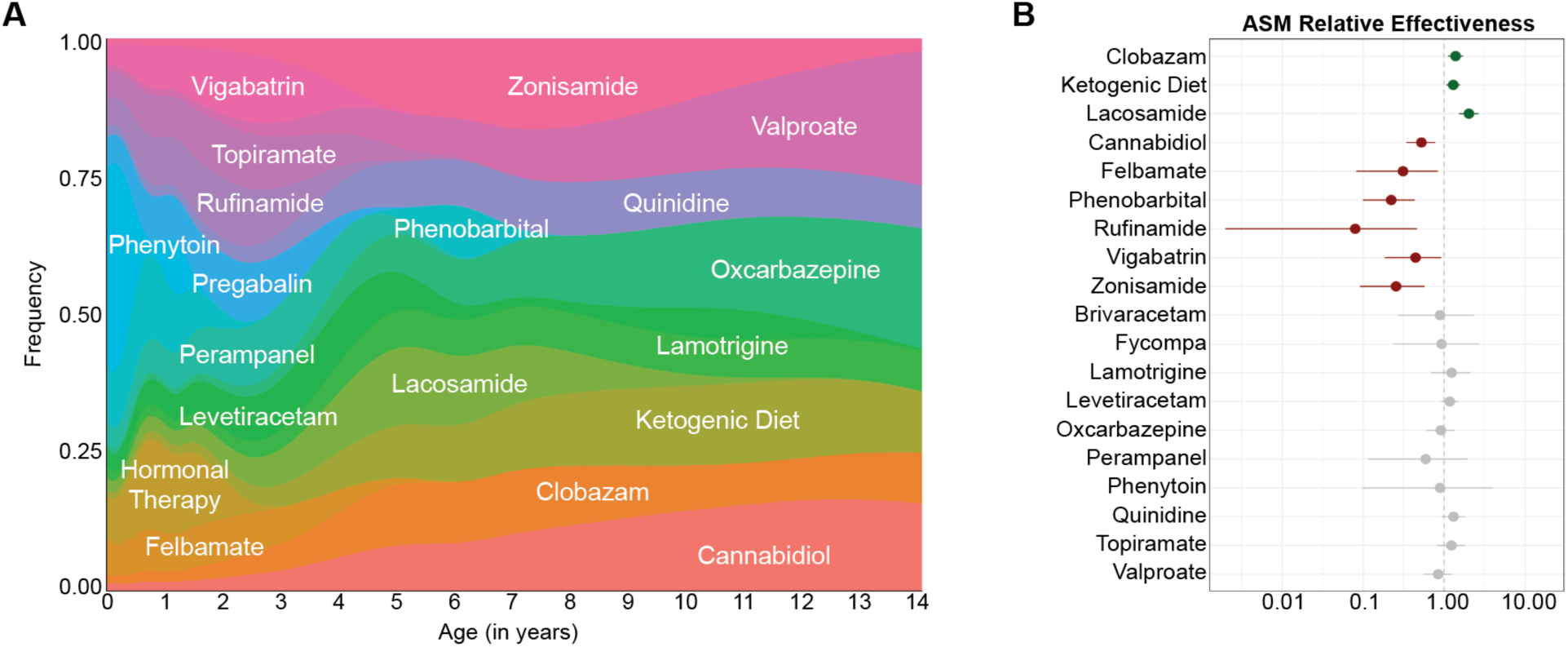
Treatment response and Anti-Seizure Medication Effectiveness. (**A**) Frequency of individuals prescribed each anti-seizure medication (ASM) over time. ASMs used by fewer than 3 individuals at any given time are not shown. (**B**) Relative effectiveness of ASMs in seizure frequency reduction or maintenance of seizure freedom. Odds ratios were calculated based on changes in seizure frequency score between consecutive monthly intervals. Green indicates a significant reduction in seizure frequency, red indicates a significant increase, and grey indicates no significant change. ASMs with insufficient clinical information are not shown.

Temporal patterns in medication prescriptions revealed a general progression of treatment strategies over the disease course. Peak use was concentrated in early infancy for several ASMs, including phenobarbital and levetiracetam at 3–4 months (*n*=31 and 28), clobazam and topiramate at 4 months (*n*=21 and 18), phenytoin and lacosamide at 3 months (*n*=14 and 7), vigabatrin at 9–10 months (*n*=11), and the ketogenic diet at 10 and 13–15 months (*n*=24). Later peak use was observed for cannabidiol at 20 months (*n*=13), quinidine from 13–21 months and at 28 months (*n*=7 and 10), felbamate at 25–26 months (*n*=7), perampanel at 26–32 months (*n*=3), valproate at 44–46 and 94–95 months (*n*=5), and zonisamide at 67 months (*n*=5). Several therapies also demonstrated lower-level sustained use across later childhood, including oxcarbazepine, levetiracetam, clobazam, the ketogenic diet, and cannabidiol. Together, this distribution reflects a pattern of evolving treatment use, highlighting the ongoing need for therapeutic adjustment over time.

To further evaluate the utility of commonly prescribed ASMs, we performed a comparative effectiveness analysis to assess ASM association with a composite clinical outcome defined as either improvement in seizure severity or maintenance of seizure freedom over a defined period (**Fig. 7B**). Two individuals did not have corresponding seizure frequencies, and one did not have recorded prescription start and end times; therefore, the final cohort consisted of 59 individuals. Clobazam (OR=1.39, 95% CI 1.12-1.72, *f*=0.85), ketogenic diet (OR=1.30, 95% CI 1.07-1.57, *f*=0.75), and lacosamide (OR=2.03, 95% CI 1.54-2.66, *f*=0.59) were each positively associated with the composite outcome of seizure improvement or sustained seizure freedom. In contrast, cannabidiol (OR=0.53, 95% CI 0.34-0.78, *f*=0.66), felbamate (OR=0.31, 95% CI 0.082-0.84, *f*=0.14), phenobarbital (OR=0.22, 95% CI 0.10-0.43, *f*=0.80), rufinamide (OR=0.080, 95% CI 0.002-0.46, *f*=0.10), vigabatrin (OR=0.45, 95% CI 0.19-0.92, *f*=0.31), and zonisamide (OR=0.24, 95% CI 0.092-0.58, *f*=0.22) were negatively associated with this outcome. Overall, these findings highlight substantial variability in medication use among individuals with *KCNT1*-related disorders, while also providing quantitative evidence that certain therapies may be more strongly associated with improved seizure outcomes than others.

### qEEG Biomarkers

To identify objective, quantifiable biomarkers capable of tracking disease progression and stratifying patients, we performed quantitative EEG analysis on 13 individuals with *KCNT1*-related disorders alongside age-matched controls (**Fig. 8A**). Random forest models, which leverage an ensemble of decision trees to recognize patterns and assign a diagnosis, trained to predict whether EEGs were from individuals with *KCNT1* or neurotypical controls performed with high accuracy (AUC = 0.906, **Fig. 8B**). Distinct spectral abnormalities drove model performance, with central alpha band power (mean decrease Gini [MDG] = 9) and parietal alpha band power (MDG = 7.5) identified as the most informative spectral features for distinguishing *KCNT1* EEGs from controls (**Fig. 8C**). Quantification of these features across developmental stages revealed age-dependent reductions in alpha and alpha/theta band power in individuals with *KCNT1*, with central alpha band power reduced across both childhood and adolescence/adulthood (*P*<0.001), parietal alpha band power reduced in childhood (*P*<0.05) and adolescence/adulthood (*P*<0.01), and alpha theta-central band power reduced in childhood (*P*<0.001) and adolescence/adulthood (*P*<0.05), relative to age-matched controls (**Fig. 8D**).

**Figure 8.**
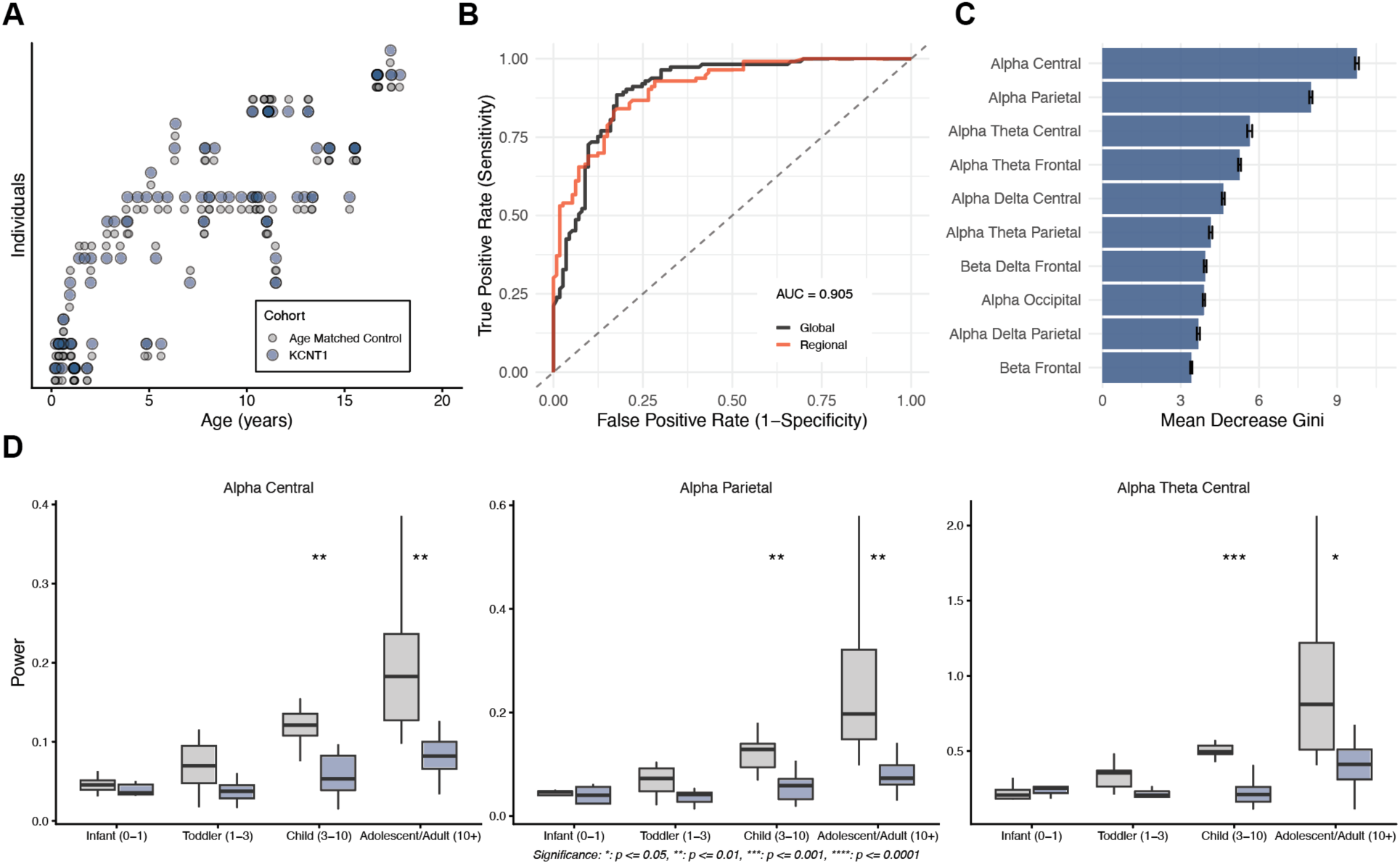
Summary of qEEG Signatures in *KCNT1.* (**A**) An inventory of EEGs collected throughout patient clinical care; each row in the *KCNT1* cohort shows EEGs for a single individual with the nearest age matched controls for each recording (sourced from unique individuals). (**B**) ROC curves demonstrating random forest model performance with global (black) and regional (red) features; AUC values were equivalent in both outcomes. (**C**) Summary of feature importance for the model using regional features. (**D**) Boxplot comparisons between age matched controls and *KCNT1* individuals (one recording randomly sampled per individual per age bin).

## Discussion

Here, we characterize the natural history of *KCNT1*-related disorders through longitudinal phenotypic reconstruction from medical records, systematic review and synthesis of published literature, and computational analyses. Together, these approaches reveal several key insights into disease progression and phenotypic variability. While overall clinical presentations exhibited identifiable trends over time within our cohort, they also underscore the complexity and heterogeneity of genotype–phenotype correlations within *KCNT1*-related disorders and clinically relevant subgroups. Our study enables detailed characterization of longitudinal seizure trajectories and developmental end points, which may inform clinical trial endpoints for the targeted therapeutic strategies under development. Notably, affected individuals had a severe and persistent seizure burden, with considerable variability in treatment approaches used to manage seizures. Despite this heterogeneity, we identified specific antiseizure medications demonstrating superior comparative effectiveness, with improvement in seizure frequency or periods of sustained seizure freedom. Lastly, we identified potential quantitative EEG spectral signatures that may provide clinically measurable biomarkers for the stratification and longitudinal monitoring of *KCNT1*-related disorders.

It has been previously reported that certain disease-causing variants in *KCNT1* can manifest across both EIDEE and ADSHE subgroups, a finding which has been confirmed in our study and reinforces the need for a comprehensive understanding of the clinical spectrum of *KCNT1*-related disorders.^3–5^ In our study of 159 individuals, genotype-phenotype analyses identified specific clinical features that were significantly associated with recurrent variants.^36^ However, variant-associated phenotypic features were predominantly aligned with EIDEE-related manifestations. In contrast, no variant-specific phenotypic features were uniquely or consistently associated with the ADSHE subgroup, highlighting limited genotype-phenotype correlations within this subgroup. Additionally, our comparison to a broader cohort of 14,893 individuals with neurodevelopmental disorders enabled confirmation of phenotypic features significantly enriched in *KCNT1*-related disorders, further contextualizing the clinical spectrum.^37,38^

Major aortopulmonary collateral artery (MAPCA), more recently described in *KCNT1*-related disorders as systemic-pulmonary collaterals (SPCs), represent a rare but potentially fatal cardiovascular phenotype exhibited in individuals with *KCNT1*-related disorders.^46,47^ We identified 7 individuals presenting with this specific phenotype, with age of diagnosis ranging from 2 to 20 months. This finding is unique due to the higher incidence observed in our cohort compared to previous reports.^46,48,49^ For instance, one study reported a prevalence of SPCs among individuals with *KCNT1*-related disorder to be 1 in 6 but described only a single confirmed case, highlighting the limited documentation and understanding of this phenotype in the literature.^48^ Our findings emphasize the importance of heightened clinical awareness and routine cardiovascular evaluations for individuals living with *KCNT1*-related disorders to facilitate early detection and management of SPCs.

Our longitudinal analysis of seizure trajectories in 62 individuals emphasizes the persistent severity of seizures across *KCNT1*-related disorders, in contrast to other neurogenetic disorders, which often exhibit greater variability in seizure severity.^32–35.42,43^ For example, individuals with *SYNGAP1*-related disorders typically develop frequent seizures later in life, usually beginning after 12 months of age (Figure 5C) .^35^ In contrast, 81% of all individuals with *KCNT1* in our study (*n*=126/156) had seizures within the first year of life. Although individuals with *SCN1A*-related disorders also present with early-onset seizures, they are generally less frequent than those seen in *KCNT1*.^42^ In our longitudinal cohort, 12 out of 21 of individuals (57%) had at least five seizures per day in the first month of life. Accordingly, *KCNT1*-related epilepsy is particularly unique with regards to its early seizure onset and consistently high seizure burden. Our EMR-based approach has allowed us to comprehensively analyze the landscape of *KCNT1*-related disorders, in addition to enhancing our ability to identify potential outcome measures to improve clinical trial readiness.^50^ Thus, the frequent and pervasive nature of seizures and developmental delays in *KCNT1*-related disorders not only underscores their clinical severity but also positions them as a reliable endpoints for future clinical trials.

The progression of developmental milestones within our cohort of 62 individuals provides a quantitative framework for assessing the extent of delay and, consequently, the severity of the developmental impact of the disorder. As described, seizures emerge early in individuals with *KCNT1*-related disorders, contributing to neurodevelopment impairment beginning in infancy. The systematic delineation of well-defined developmental end points, an aspect not comprehensively addressed in prior studies, strengthens clinical trial readiness by enabling meaningful outcome measurement and supporting the evaluation of therapeutic efficacy beyond seizure reduction alone. Collectively, these findings position developmental milestone trajectories in *KCNT1*-related disorders as critical endpoints for capturing disease progression and guiding future therapeutic intervention strategies.

Our analysis of medication prescription data over time provides insights into the specific timeline of treatments among a cohort of individuals with *KCNT1*-related disorders. Studies investigating quinidine, cannabidiol (CBD), and the ketogenic diet have identified these as beneficial therapies for managing *KCNT1*-related disorders, with evidence indicating that quinidine and the ketogenic diet are more effective than conventional ASMs.^39,40^ Building on these findings, our analysis revealed that quinidine was not significantly associated with either seizure reduction or maintenance of seizure freedom, whereas the ketogenic diet was significantly associated with both outcomes. In contrast to previous reports, CBD was negatively associated with both seizure freedom maintenance and seizure frequency reduction, highlighting variability in treatment response across individuals. Notably, 58 out of 62 individuals were prescribed levetiracetam at some point during their lifetime, most commonly within the first four years of life; however, despite its widespread use, levetiracetam was not significantly associated with either superior seizure frequency reduction or maintenance of seizure freedom. This finding may, in part, reflect its widespread use as a first-line anti-seizure medication in pediatric epilepsy, raising the possibility that observed effectiveness is influenced by treatment prevalence and duration rather than drug-specific efficacy alone.^51,52^ Taken together, these results underscore the heterogeneity in medication strategies for *KCNT1*-related disorders; nevertheless, our analyses identify medications associated with differential seizure outcomes, providing important insight into real-world treatment effectiveness and informing future therapeutic strategies.

Quantitative EEG analysis, while preliminary, further supports the emerging role of EEG spectral features as objective biomarkers in genetic epilepsies. The developmental persistence of these abnormalities suggests that *KCNT1*-related disorders may involve persistent disruptions in cortical network organization and maturation, rather than isolated electrophysiologic abnormalities alone. These findings align with prior work demonstrating that qEEG-derived spectral signatures can differentiate genetic epilepsies and predict neurologic outcomes, reinforcing the broader utility of quantitative electrophysiologic biomarkers for disease stratification and longitudinal tracking.^40,52^ Within *KCNT1*-related disorders, the integration of longitudinal real-world EEG recordings with machine learning approaches highlights the potential for scalable, non-invasive biomarkers capable of supporting disease stratification, tracking progression across development, and informing future biomarker-driven clinical trials.

Our study leverages real-world data derived from EMRs across multiple sources, including CHOP and Citizen Health, which introduces several inherent limitations. First, as is common in EMR-based analyses, data completeness is variable and dependent on the clinical context of individual encounters, resulting in missing or inconsistently documented features. Importantly, the absence of a recorded phenotype does not necessarily indicate true absence; rather, it may reflect incomplete clinical documentation. Second, the integration of data from multiple sources introduces heterogeneity in data extraction practices and phenotypic annotation, which may contribute to variability across the cohort despite efforts to standardize features through mapping to HPO terms. In addition, seizure burden was assessed using the PEHLS seizure frequency scale, which captures broad categorical severity rather than exact seizure counts. Because the highest category includes all individuals with more than five seizures per day, this scale introduces a ceiling effect that may obscure clinically meaningful changes among individuals with very high seizure burdens. For example, an individual whose seizure frequency decreases from dozens of seizures per day to fewer, but still more than five per day, would retain the same PEHLS score despite meaningful clinical improvement. Finally, our longitudinal data exhibit an uneven temporal distribution, with earlier life stages more densely represented and comparatively fewer observations captured at older ages. This imbalance likely reflects both the structure of clinical follow-up patterns and the nature of real-world data collection and may limit the ability to fully characterize phenotypic trajectories across the lifespan.

Additionally, our study includes analyses across two cohorts: a larger cohort of 159 individuals, incorporating both literature-reported cases and longitudinal data, and a subset of 62 individuals with longitudinal EMR-derived phenotyping. While the larger cohort enabled comparisons of recurrent variants, subgroup sizes remained limited, with the largest recurrent variant group represented by only 23 individuals. This constraint should be considered when interpreting genotype-phenotype associations, as limited subgroup sizes may reduce statistical power and stability of observed associations. Comparisons across clinical subgroups, EIDEE and ADSHE, were similarly imbalanced, with substantially fewer individuals in the ADSHE group (*n*=22) compared to EIDEE (*n*=132), further limiting the generalizability of subgroup-specific findings. Nevertheless, these analyses provide meaningful insight into temporal patterns of disease progression, supporting a more nuanced, longitudinal understanding of *KCNT1*-related disorders that may inform future clinical and therapeutic strategies.

In sum, by integrating longitudinal phenotypic reconstruction from real-world medical records with systematic literature synthesis and computational analyses, we delineate the natural history and heterogeneity of *KCNT1*-related disorders while identifying shared longitudinal trends and clinically meaningful subgroup differences. Seizure burden was severe and sustained despite diverse treatment strategies, supporting persistently high seizure frequencies, alongside key developmental end points, as clinically relevant outcomes to capture disease severity and therapeutic impact in future trials. Leveraging real-world data to map disease trajectories enables pragmatic, measurable outcome selection to accelerate trial design and evaluation as targeted therapies advance.

## Data Availability

All data produced in the present study are available upon reasonable request to the authors.

## Acknowledgements

We thank the *KCNT1* Epilepsy Foundation, both by funding this work through a seed grant to JLM and for their ongoing consultation and feedback on this project. Special thanks are due to Ali Rosenberg, Brad Bryan, and Sarah Drislane. We would also like to acknowledge all the patients and families who have contributed data through participation in research and by sharing medical records with Citizen Health.

## Abbreviations

ACMG: American College of Medical Genetics and Genomics
ASM: anti-seizure medications
DEE: developmental and epileptic encephalopathy
EOEE: early onset epileptic encephalopathy
EIDEE: early infantile developmental and epileptic encephalopathy
ADSHE: autosomal dominant sleep-related hypermotor epilepsy
HPO: Human Phenotype Ontology
NDD: neurodevelopmental disorders
qEEG: quantitative electroencephalogram

